# Feasibility of AI-Guided Handheld Echocardiography for Rheumatic Heart Disease Screening by Non-Cardiologists in Rural Nepal: A Mixed-Methods Diagnostic Accuracy Study

**DOI:** 10.64898/2026.11.12.26367974

**Authors:** Husameddin Alattar, Dhyana Magesh, Angela Pal, Rache Mureau-Haines, Taylor Young, Keshet Ronen, David Zhang, Jay D Pal

## Abstract

**Background:** Rheumatic heart disease remains a major cause of preventable morbidity and mortality in rural Nepal, where access to cardiovascular specialists and echocardiography is limited. We evaluated the diagnostic accuracy and feasibility of AI-guided echocardiography performed by nurse practitioners for community-based RHD screening.

**Methods:** We conducted a triangulation mixed-methods study (QUAN > qual) combining a paired cross-sectional diagnostic accuracy study with semi-structured interviews during community screening camps in Damak, Nepal, in November 2025. Nurse practitioners performed AI-guided handheld echocardiography using EchoNous Kosmos Plus devices, with cardiologist overread serving as the comparator. Diagnostic performance was assessed using sensitivity, specificity, and kappa. Interviews were analyzed thematically.

**Results:** Of 90 participants enrolled, 76 were included in the final analysis. Mean age was 50.8 years, 57.8% were male, and 42.2% had undergone prior cardiac evaluation. Detection of any valvular lesion demonstrated 95.8% sensitivity and 100% specificity, with near-perfect agreement with cardiologist interpretation (κ=0.97). Lesion-specific performance included mitral regurgitation sensitivity 66.7% and specificity 97.3% (κ=0.55), mitral stenosis sensitivity 100% and specificity 97.2% (κ=0.81), aortic regurgitation sensitivity 90.9% and specificity 96.9% (κ=0.84), and tricuspid regurgitation sensitivity and specificity of 100% (κ=1.00). Mean scan duration was 5.8 minutes. Portability, AI-assisted acquisition, and remote review facilitated implementation, while training needs, workflow constraints, and technical issues were barriers.

**Conclusions:** AI-guided handheld echocardiography performed by nurses demonstrated high diagnostic accuracy and feasibility for rural RHD screening. AI-assisted task-shifting may expand access to early RHD detection in underserved settings.

## Introduction

Rheumatic heart disease (RHD) remains a leading cause of preventable morbidity and mortality in rural Nepal.^1^ RHD is a chronic sequela of acute rheumatic fever (ARF), an immune-mediated inflammatory disease that typically follows infection with group A Streptococcus and occurs most commonly in children and adolescents.^2^ Through molecular mimicry, streptococcal antigens can trigger cross-reactive immune responses against host tissues, including cardiac structures.^2^ Recurrent episodes of ARF drive a destructive cycle of cumulative valvular inflammation, scarring, and fibrosis, culminating in permanent structural remodeling of the heart valves.^2^ This progressive damage often results in hemodynamically significant lesions, such as mitral or aortic stenosis and regurgitation, which severely compromise cardiac function and lead to debilitating, life-limiting cardiovascular complications.^3^

Secondary prophylaxis with regular penicillin injections remains the gold-standard, cost-effective strategy for preventing recurrent ARF and limiting progression of RHD. However, many patients are not diagnosed until symptomatic, when advanced disease has developed, thus missing the opportunity for early intervention when prophylaxis may provide the greatest benefit.^4^ This has made early detection of asymptomatic or subclinical RHD an important prevention strategy.

Traditional approaches relied on clinical examinations and cardiac auscultation; however, these methods have limited sensitivity, often failing to detect subclinical disease.^4^ In the past decade, echocardiography has evolved to identify RHD more accurately, including disease without a clinically detectable murmur.^4^ Advances in portable echocardiography have further expanded the feasibility of screening in remote and resource-limited settings, creating opportunities to detect disease earlier, initiate secondary prophylaxis, and potentially reduce RHD-related morbidity and mortality.^5^

This shift toward echocardiographic screening has been accompanied by parallel efforts to standardize how RHD is diagnosed. In 2023, the World Heart Federation (WHF) released updated guidelines for the echocardiographic diagnosis of RHD, revising the widely used 2012 criteria to reflect a decade of accumulated evidence and end-user feedback.^6^ The updated guidelines refine the screening and confirmatory criteria used for active case finding and introduce a stage-based classification that stratifies patients according to their risk of progression to advanced valvular disease.^6^ By standardizing minimum diagnostic criteria for use in RHD-endemic, resource-limited settings, the 2023 guidelines aim to support earlier, more consistent detection and timely initiation of secondary prophylaxis.

Building on this diagnostic framework, the World Health Organization (WHO) released its first guideline on the prevention and diagnosis of ARF and RHD in 2024, developed in partnership with the WHF. The guideline consolidates recommendations spanning primary and secondary prevention, diagnostic pathways, and clinical management, providing a single evidence-based reference intended for use across primary, district, and referral levels of care in RHD-endemic countries.^7^ It responds to a gap identified by WHO itself: despite the 2018 World Health Assembly resolution calling for a coordinated global response to ARF/RHD, countries lacked a unified technical resource to guide implementation. By translating the 2023 WHF echocardiographic criteria and other recent evidence into actionable, health-system-level guidance, the 2024 WHO guideline aims to support countries in strengthening screening, prophylaxis, and referral systems for RHD.^7^

Nepal has undertaken several initiatives to strengthen RHD prevention and care, but important gaps remain. In 2007, the Nepal Heart Foundation, with government support, launched a comprehensive RHD control program incorporating community awareness, free medication, patient registries, health worker training, clinical guidelines, and secondary prophylaxis.^8^ However, inadequate funding led to discontinuation of the program and a shift toward subsidizing heart valve surgery, weakening systems for secondary prevention.^8^ Consequently, adherence to recommended prophylaxis remains limited. One study found that only 21.9% of patients received intramuscular benzathine penicillin, while most received oral penicillin instead.^9^

These challenges are compounded by substantial workforce limitations, particularly in specialized cardiovascular care. Nepal has fewer physicians per capita than the global average and a critical shortage of cardiac surgeons, with approximately 25 surgeons reported nationally by 2022,^10^ giving Nepal one of the lowest per-capita capacities in South Asia. Although cardiac surgical capacity has expanded to approximately 15 centers performing around 2,000 procedures annually, services remain concentrated in the Kathmandu Valley and insufficient to meet national demand.^11^

More recently, Nepal has pursued decentralization through the PEN-Plus initiative, launched in 2021 to strengthen management of severe noncommunicable diseases, including RHD, at district-level facilities.^12^ Implemented through the Kathmandu Institute of Child Health, the initiative emphasizes training local health workers, expanding access to diagnostic tools, and strengthening referral pathways between district and tertiary centers.^12^ These efforts represent important progress toward equitable RHD care; however, persistent shortages of specialized personnel, limited secondary prophylaxis systems, and geographic concentration of advanced services underscore the need for scalable approaches that enable earlier RHD detection and decentralized screening in underserved communities.

AI-guided handheld echocardiography devices offer a promising approach to addressing persistent barriers to RHD screening in resource-limited settings by enabling non-cardiologist healthcare workers with limited echocardiographic training to acquire clinically interpretable cardiac images. By expanding screening capacity beyond specialized cardiovascular centers, this technology may facilitate earlier identification of previously undiagnosed disease and timely initiation of secondary prophylaxis, potentially reducing progression to advanced valvular damage. However, evidence regarding the diagnostic performance and real-world feasibility of AI-guided echocardiography when used by non-cardiologists in low-resource settings remains limited.

### Study Objectives

The primary objective of this study was to determine the diagnostic accuracy of AI-guided handheld echocardiographic screening for rheumatic heart disease when performed by non-cardiologist providers in a rural, resource-limited setting in Nepal. Specifically, we aimed to estimate the sensitivity, specificity, and Cohen’s kappa agreement coefficient of screening examinations performed by nurse practitioners using the EchoNous Kosmos system, with cardiologist overread as the reference standard, both overall and for individual valve lesions.

Secondary objectives were to assess the quality and ease of image acquisition by non-cardiologist screeners, and to characterize, through semi-structured interviews with field providers, the perceived facilitators, barriers, and recommendations for implementing AI-guided echocardiographic screening in routine practice.

## Methods

### Study Design

This study employed a triangulation mixed-methods design with a quantitative-dominant approach (QUAN > qual). The quantitative component consisted of a prospective paired cross-sectional diagnostic accuracy study comparing handheld echocardiographic screening performed by non-cardiologist providers using an AI-guided ultrasound system vs. cardiologist overread interpretation of the saved studies. The qualitative component consisted of semi-structured interviews with the providers who performed the screening studies to explore implementation feasibility, usability, workflow integration, and barriers and facilitators to deployment in rural settings.

### Training of Non-Cardiologist Screeners

The screening team consisted of two nurse practitioners (NPs) who performed the AI-guided echocardiographic examinations and one cardiologist who conducted expert overread interpretation. Neither NP had prior experience performing formal echocardiographic examinations or screening for rheumatic or valvular heart disease. Although both were familiar with point-of-care ultrasound (POCUS) applications in clinical practice, neither routinely performed POCUS examinations. The interpreting cardiologist is board-certified in echocardiography in the United States. Before initiation of the screening camps, both NPs completed a one-day hands-on training conducted by the device manufacturer. Training covered device operation, acquisition of standard cardiac views, use of AI-guided image acquisition features, and study-specific screening procedures.

### Screening Camps and Screening Device

Community-based screening camps were conducted over a two-week period in November 2025 in Damak, a rural agricultural municipality in Jhapa District, Nepal. Screening camp sites were selected based on existing epidemiological data on RHD prevalence, prioritizing districts with the highest estimated disease burden to maximize case detection and study yield. Community engagement and recruitment for each camp were conducted through localized advertising via print media and radio broadcasts within the selected district in the weeks preceding the event, in order to inform residents of the screening date, location, and eligibility. On the day of the camp, all participants underwent a standardized registration process to collect demographic and identifying information, followed by a structured history and physical examination performed by trained clinical staff, with particular attention to symptoms and signs suggestive of rheumatic heart disease. Vital signs and a 6-lead ECG were obtained for every participant as part of the standard screening protocol. Echocardiographic screening was performed on all participants regardless of clinical or ECG findings. This was to capture the full spectrum of RHD, including subclinical or latent disease that might otherwise go undetected by history, exam, or ECG alone, and to generate an accurate estimate of true disease prevalence within the screened population.

Echocardiographic screenings were performed using the EchoNous Kosmos Plus handheld ultrasound device, shown in Figure 1, with integrated real-time AI-guided image acquisition support, compatible with the iOS operating system and paired with a 13-inch Apple iPad. The Kosmos software is unique in providing real-time guidance on image acquisition by directing the user on how to position the probe via on-screen animations, grading of image quality via on-screen scoring bars, and automated labeling of anatomic structures. In addition AI-determined ejection fraction was documented.

**Figure 1.**
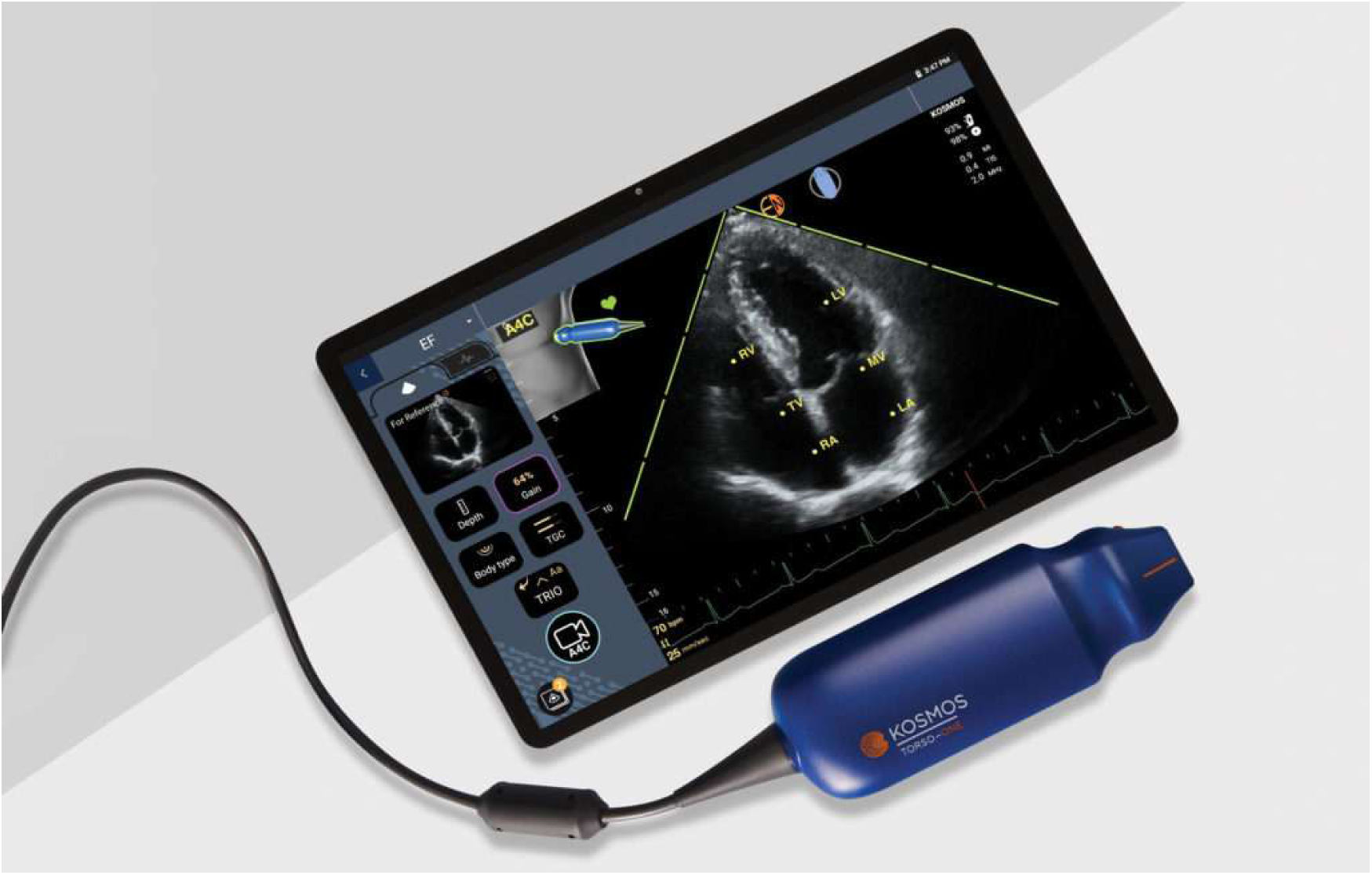
EchoNous Kosmos AI-assisted echocardiography probe attached to a display device.

Echocardiographic screening was performed using a standardized, abbreviated protocol consisting of four views: parasternal long-axis, parasternal short-axis, apical four-chamber, and apical five-chamber. This focused imaging protocol was designed to allow rapid assessment of valvular structure and function, chamber size, and hemodynamically significant regurgitation or stenosis in a field setting, while maintaining sufficient diagnostic yield to identify morphologic and functional features consistent with rheumatic heart disease. For screening purposes, participants with any detected structural valvular abnormality were classified as screen-positive and referred for confirmatory diagnostic evaluation.

### Participants

A total of 90 participants were recruited during the screening camps. Eligible participants were individuals attending the screening camps who were able to undergo handheld echocardiographic examination and provide informed consent or assent. Participants were excluded if they had hemodynamic instability or other conditions that precluded safe echocardiographic assessment.

### Quantitative Statistical Analysis

Descriptive statistics were used to summarize participant characteristics, including age, gender, medical history, and scan duration. Diagnostic accuracy analyses compared AI-guided echocardiography by non-specialist providers vs. cardiologist overread as the gold standard. We used 2 × 2 contingency tables to calculate true positives, true negatives, false positives, false negatives, sensitivity, and specificity. Inter-rater agreement between non-cardiologist screeners and cardiologist interpretation was assessed using Cohen’s kappa statistic.

For the “any valvular lesion” category, each participant was counted only once, regardless of the number of valvular lesions detected. In lesion-specific analyses, participants with multiple lesions were included in each applicable lesion category; therefore, lesion-specific counts are not mutually exclusive and may exceed the number of participants with any valvular lesion.

A paired-samples t-test was used to compare mean ejection fraction (EF) measurements obtained by the nurse practitioner and the cardiologist.

Secondary quantitative outcomes included NPs’ mean confidence in image quality score using a 3-point scale (1 = poor, 2 = adequate/limited, 3 = good); mean ease of image acquisition score using a 3-point scale (1 = low, 2 = moderate, 3 = high); and mean scan duration, defined as the time in minutes from probe placement to completion of image acquisition. We used SPSS and Microsoft Excel software for quantitative analysis.

### Qualitative Data Collection and Analysis

All screening providers involved in the screening camps (two nurse practitioners and one cardiologist) participated in the qualitative component. Semi-structured interviews lasting approximately 30–45 minutes were conducted and audio-recorded with participant consent. Interviews were conducted by video call. Verbatim transcripts were generated. Thematic saturation was not the goal of this study given the small, purposive, whole-team sample.

Interview domains included training adequacy, usability of the handheld ultrasound device and AI guidance system, image acquisition experience, workflow and scan duration, applicability in routine clinical practice, environmental constraints (including space, electricity, and internet connectivity), cultural acceptability, data management considerations, and recommendations for improvement.

Interview transcripts were independently coded by two researchers (HA and AP), and discrepancies were resolved by discussion between the two coders before thematic analysis was conducted. Coding and qualitative data management were conducted using Dedoose (SocioCultural Research Consultants, LLC, Redondo Beach, California) and Microsoft Word software (Microsoft Corporation, Redmond, Washington).

### Ethics approval

IRB approval was obtained from the University of Washington (IDs STUDY00024715 and STUDY00019788) and the Social Welfare Council of Nepal. All participants provided verbal informed consent for data collection. Written documentation of consent was waived.

## Results

### Quantitative results

#### Participant characteristics

Ninety participants were registered in the screening camps between November 1 and November 15, 2025, in Damak, Nepal. Table 1 presents participants’ demographic and medical characteristics. Due to patient factors, lack of consent, and workflow issues, only 76 patients underwent echocardiographic screening using the EchoNous Kosmos devices by the nurse practitioners. Participants had a mean age of 50.2 years (range, 19–86 years), and 42.2% were female. Overall, 7.7% were diabetic, 12.2% had hypertension, 6.6% had a history of coronary artery disease, 1.1% were diagnosed with heart failure, and only 42.2% had ever had a cardiovascular evaluation.

**Table 1.** Campaign participants’ demographic and medical characteristics.

|  | Registered Cohort<br>Mean (SD) or n (%)<br>N = 90 | Screened Cohort<br>Mean (SD) or n (%)<br>N = 76 |
|---|---|---|
| Age | 50.8 | 50.2 |
| Female Gender | 38 (42.2%) | 34 (44.7%) |
| Diabetes | 7 (7.7%) | 7 (9.2%) |
| HTN | 11 (12.2%) | 9 (11.8%) |
| Heart failure | 1 (1.1%) | 1 (1.3%) |
| History of coronary artery disease | 6 (6.6%) | 6 (7.8%) |
| History of prior cardiac evaluation | 38 (42.2%) | 33 (43.4%) |

Table 2 summarizes the overall and the individual valve lesion diagnostic performance results of NPs compared to the reference standard. Figure 2 shows the comparison between NPs and reference standard diagnostic performance. Across the screened cohort, the prevalence of any lesion per reference test was 31.6 cases per 100 patients. Individual lesion prevalence was 3.9 per 100 for mitral regurgitation (MR), 6.6 per 100 for mitral stenosis (MS), 14.5 per 100 for aortic regurgitation (AR), 13.2 per 100 for tricuspid regurgitation (TR), 1.3 per 100 for pulmonary regurgitation (PR), and no cases were found for aortic stenosis (AS).

**Figure 2.**
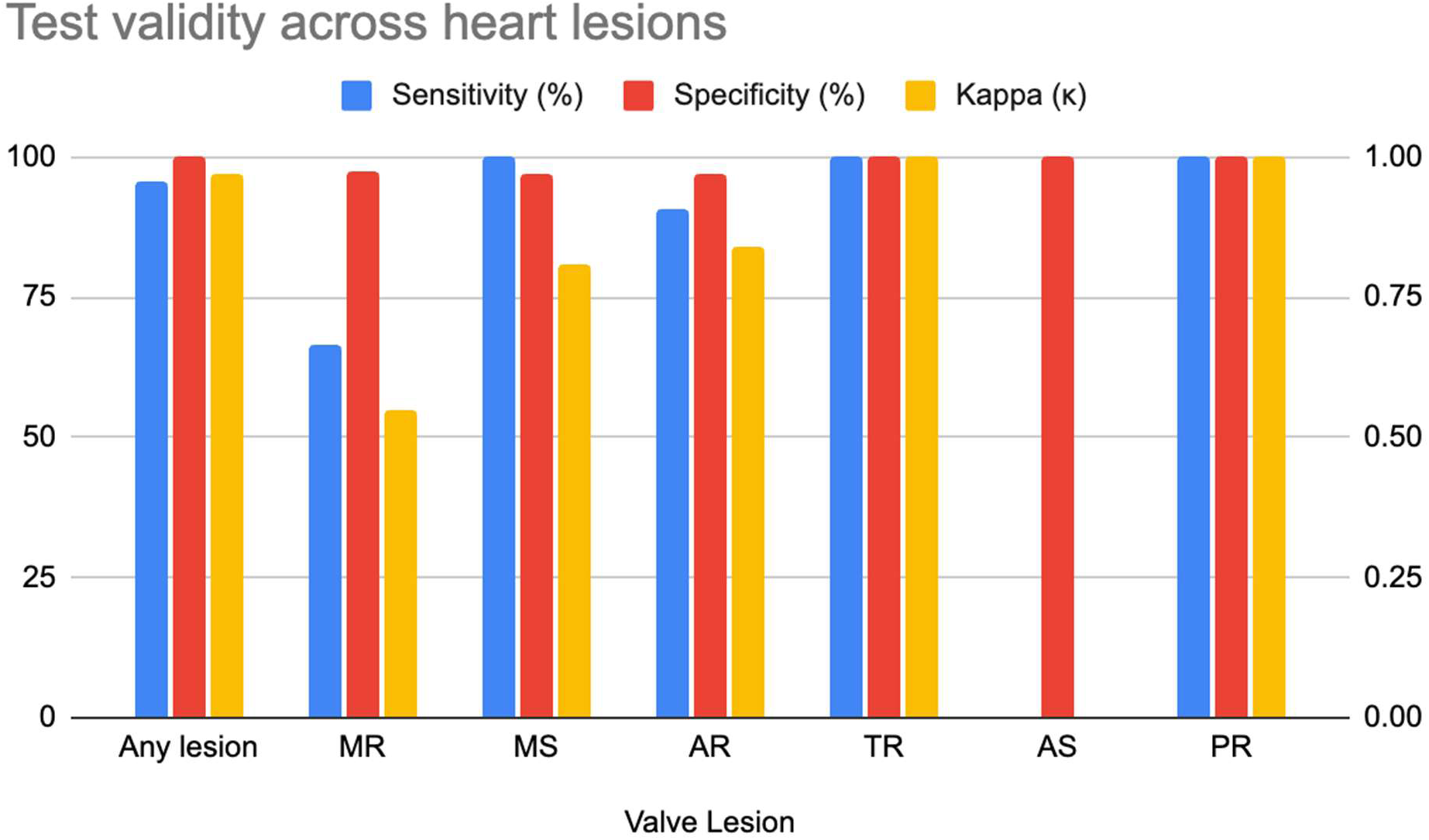
Display of NPs’ diagnostic performance across individual valve lesions

**Table 2.** Overall and individual valve lesion diagnostic performance of NPs*.

| **Valve lesion** | **Sensitivity (%)** | **Specificity (%)** | **Kappa** | **# of true positive cases** | **Agreement Interpretation** |
|---|---|---|---|---|---|
| **Any lesion** | 95.8 | 100 | 0.97 | 23 | Almost perfect |
| **MR** | 66.7 | 97.3 | 0.55 | 2 | Moderate |
| **MS** | 100 | 97.2 | 0.81 | 5 | Almost perfect |
| **AR** | 90.9 | 96.9 | 0.84 | 10 | Almost perfect |
| **TR** | 100 | 100 | 1 | 10 | Perfect |
| **AS** |  | 100 |  | 0 | Not estimable |
| **PR** | 100 | 100 |  | 1 | Not estimable |

\*Lesion-specific categories are not mutually exclusive. Participants with multiple lesions may be represented in more than one category.

NPs were able to detect 23 out of the 24 positive cases, demonstrating high diagnostic accuracy, with a sensitivity of 95.8% and specificity of 100%, and almost perfect agreement with the reference standard (kappa = 0.97).

Across individual valve lesions, performance varied. For MR, sensitivity was lower at 66.7%, while specificity remained high at 97.3%, with moderate agreement (kappa = 0.55). In contrast, MS showed excellent performance, with 100% sensitivity, 97.2% specificity, and almost perfect agreement (kappa = 0.81). Similarly, AR demonstrated strong performance, with 90.9% sensitivity, 96.9% specificity, and almost perfect agreement (kappa = 0.84). For TR, NPs achieved 100% sensitivity and specificity, with perfect agreement (kappa = 1.00). However, all these results should be interpreted cautiously due to the relatively small number of positive cases. Our dataset recorded one case of PR, yielding 100% sensitivity and specificity (kappa = 1.00) for NP detection, which should also be interpreted with caution. Analysis for AS could not be performed due to the absence of positive cases in the dataset.

Additionally, comparison of EF measurements between NPs and the reference standard using a paired t-test (n = 46 complete pairs) found no significant difference between methods (mean difference 2.50%, 95% confidence interval 0.56 to 5.56). Out of a maximum score of 3 points, nurse practitioners reported an average ease of image acquisition score of 2.36 and an average confidence in image-quality score of 2.69. The mean examination time per patient was 5.8 minutes, with a range of 2–20 minutes.

### Qualitative results

#### Theme 1: Capacity Building and User Competence

Participants consistently emphasized that successful adoption of AI-supported echo depended on adequate preparation, repeated practice, and the ability to interpret AI guidance in light of clinical judgment. While the technology reduced some technical barriers to image acquisition, participants described competency as developing progressively through hands-on experience rather than through training alone.

Although participants generally viewed the initial training positively, they noted that additional practice was necessary to become comfortable with both image acquisition and the device interface. One nurse practitioner described supplementing the formal training with independent practice sessions using family members and volunteers, explaining:

> *“I definitely had to practice both the exam and using the interface… I practiced on my own with my family and some volunteers”* (Interviewee 1).

Participants reported that confidence and efficiency improved substantially with repeated use in the field, particularly when encountering patients with varying body habitus and clinical presentations. As one participant noted:

> *“The more time I spent screening, the faster and more efficient that I got”* (Interviewee 2).

Despite feeling adequately prepared to begin screening, participants suggested that additional structured training materials, supervised practice, and ongoing mentorship would further strengthen user preparedness.

Participants also described a gradual process of building trust in the AI-assisted platform. While they expressed confidence in their ability to obtain diagnostic-quality images, they were often more cautious about relying on AI-generated assessments. One participant explained:

> *“I had very high confidence in my ability to gather good clinical images, but sometimes I didn’t trust the AI’s labeling of whether an image was good or not”* (Interviewee 1).

Rather than accepting AI outputs uncritically, participants frequently compared AI feedback against their own assessment and sought confirmation from the supervising cardiologist when uncertainty arose. Repeated exposure to screening appeared to increase their confidence in identifying abnormal findings, but trust in the technology remained conditional and closely tied to users’ clinical experience.

Across interviews, participants consistently framed AI as an assistive tool rather than a replacement for clinical expertise. Clinical judgment, prior exposure to echo, and specialist consultation remained central to screening and referral decisions. One nurse practitioner described their approach to uncertain findings:

> *“If there was anything questionable, I would get the highest quality imaging possible and then refer to the cardiologist”* (Interviewee 2).

Similarly, another participant emphasized that while AI improved image acquisition, it did not meaningfully influence clinical decision-making, stating:

> *“AI was helpful in acquiring images and getting good image acquisition… but it was not useful in clinical decision-making”* (Interviewee 1).

Collectively, these findings suggest that while AI-supported echo may facilitate task-shifting of RHD screening to non-cardiologists, successful implementation will require structured training pathways, continued opportunities for skill development, and clearly defined mechanisms for specialist referral.

#### Theme 2: AI guidance improved image acquisition and usability, although technical and diagnostic limitations remained

Participants generally viewed the AI-enabled devices as user-friendly and valuable for supporting image acquisition by non-specialists. The AI-guided interface was particularly helpful for novice users, providing real-time feedback on probe positioning and image quality. As one participant explained:

> *“Especially if you were like a novice…the AI system could be very, very helpful… telling you, like, ‘Oh, this is a good quality image’”* (Interviewee 2).

Similarly, the cardiologist emphasized the importance of AI support for nurses and other non-specialist screeners, noting:

> *“They don’t do this every day. So the AI can help them”* (Interviewee 3).

Participants reported that these features increased their confidence in obtaining clinically useful images and reduced some of the technical barriers traditionally associated with echocardiography.

Despite these advantages, participants also identified several technological limitations. Intermittent hardware and software issues, including overheating, system errors, and probe disconnections, occasionally disrupted workflow. One participant recalled:

> *“The system overheated once or twice… and then a couple times my setup had errors… so I had to restart the system”* (Interviewee 1),

while another described occasional probe disconnections that required restarting the platform (Interviewee 2). Participants also reported that AI-generated feedback was sometimes inconsistent, particularly when evaluating abnormal studies. In some instances, the AI classified high-quality images as suboptimal or failed to capture clinically relevant views. As one participant explained:

> *“Sometimes the AI feedback would be helpful, but… you would have really good views… and it would say, ‘Oh, this view is suboptimal’”* (Interviewee 1).

Another described the AI grading system as *“a little glitchy and not as seamless as what it could have been”* (Interviewee 2).

While participants acknowledged these limitations, they generally agreed that the technology was sufficient for screening purposes. However, they emphasized that the devices were not intended to replace comprehensive diagnostic echo or specialist interpretation. As the cardiologist noted:

> *“They are not designed for more accurate calculation… of valve severity… But… to screen for our purpose, I do think that’s good enough for us”* (Interviewee 3).

Overall, participants viewed the technology as a practical and effective screening tool that improved image acquisition and usability, while recognizing that technical limitations and diagnostic uncertainty still necessitated clinical oversight and specialist review.

#### Theme 3: AI-supported echocardiography shows strong potential for rural RHD screening, but successful implementation depends on addressing workforce, workflow, and infrastructure challenges

Participants viewed AI-supported echocardiography as a promising approach for expanding RHD screening in rural and underserved communities, citing the device’s portability, ease of deployment, and potential to connect non-specialist providers with remote cardiology expertise. The technology was perceived as particularly well suited for resource-constrained environments, with one participant noting:

> *“It worked very well in an austere environment… there was not consistent electricity… not air conditioning… not Wi-Fi available, but the system still worked very well” (Interviewee 1).*

Portability emerged as a major advantage, as participants described the device as compact and easy to transport to remote settings. As one nurse practitioner explained:

> *“One of the things that is really, really nice about it is its portability… you can take it wherever you physically go” (Interviewee 2)*.

Participants also felt that image quality was sufficient for real-world screening and, in some cases, comparable to imaging obtained in hospital settings. Combined with AI-guided acquisition and the ability to share images remotely, participants believed the technology could substantially expand access to cardiac screening in areas with limited specialist availability. One participant highlighted this potential, stating:

> *“There’s promise… to have people who aren’t formally trained… get imaging remotely and send it to another cardiology team to review” (Interviewee 2)*.

Despite this optimism, participants identified several barriers that could affect implementation and scale-up. Human resource limitations and the need for ongoing training were frequently cited concerns. One participant noted:

> *“Human resources are going to be a barrier, like getting people trained and comfortable with the system” (Interviewee 1)*.

Workflow challenges were also common, including language barriers, high patient volumes, and maintaining efficient screening processes without compromising diagnostic quality. As one participant explained:

> *“We have too many patients… You want to screen more patients, but you don’t want to shorten your protocol. If you shorten too much, you’re going to miss some of the disease” (Interviewee 3).*

Environmental constraints such as heat, background noise, limited space, and privacy concerns further complicated field implementation.

Participants nevertheless believed the technology could be scaled successfully if these operational challenges were addressed. Key requirements for broader implementation included hands-on training with real patients, improved internet connectivity to facilitate image sharing, and continued evaluation of emerging AI-enabled devices. As one participant explained:

> *“If you solve these two, the device can be widely used… train the staff with the patient… and good Wi-Fi data can be shared” (Interviewee 3)*.

Cost was also viewed as a facilitator, with handheld AI-enabled devices perceived as substantially less expensive than traditional echocardiography systems. Overall, participants viewed AI-supported echocardiography as a scalable and potentially transformative tool for rural RHD screening, but emphasized that successful implementation would require investments in training, infrastructure, workflow optimization, and healthcare system capacity.

In summary, participants viewed AI-supported echocardiography as a feasible and promising approach for expanding RHD screening through task-shifting to non-cardiologists. The devices were considered portable, generally user-friendly, and adequate for screening in rural field settings. However, participants emphasized that AI should be understood as a supportive tool for image acquisition rather than a replacement for clinical judgment or specialist interpretation.

Successful implementation will require structured training, reliable workflows, local adaptation, referral pathways, and continued cardiology oversight.

## Discussion

This mixed-methods study demonstrates that AI-guided handheld echocardiography performed by non-cardiologist providers is both diagnostically accurate and operationally feasible for community-based RHD screening in rural Nepal. Nurse practitioners achieved high overall sensitivity and excellent specificity for detecting valvular abnormalities, with almost perfect agreement with cardiologist overread for identifying any valvular lesion. Although diagnostic performance varied across individual valve lesions, the technology consistently demonstrated sufficient accuracy to support community screening. However, these results should be interpreted cautiously given the small number of valvular lesions detected in the study population. The qualitative findings complemented these results by demonstrating that providers viewed the devices as portable, intuitive, and supportive of image acquisition while recognizing the continued need for structured training and clinical judgment. Together, these findings suggest that AI-assisted handheld echocardiography represents a promising task-shifting strategy for expanding access to RHD screening in resource-limited settings.

The high diagnostic performance observed in this study is encouraging given the limited training received by the nurse practitioners and the challenging environment in which screening was performed. Overall specificity was particularly high, suggesting that nurse practitioners were able to reliably identify normal examinations and minimize unnecessary referrals. One possible explanation is that both the AI guidance system and providers were repeatedly exposed to normal cardiac anatomy during training and screening, allowing deviations from typical anatomy to be more readily recognized and referred for further evaluation. In contrast, sensitivity was comparatively lower for mitral regurgitation than for other lesions. Mild regurgitant lesions may be inherently more difficult to visualize using handheld ultrasound because of their dependence on image quality, Doppler optimization, and subtle hemodynamic findings. Although the reasons for this difference remain uncertain, it highlights an important area for future refinement of both AI algorithms and user training. Agreement in ejection fraction measurements between nurse practitioners and cardiologist interpretation was also high, likely reflecting the automated measurement algorithms incorporated into modern handheld ultrasound systems that use computational methods similar to those of conventional echocardiography. Clinically, these findings suggest that AI-guided handheld devices can provide sufficiently reliable screening when operated by trained non-cardiologists, while emphasizing that positive or equivocal findings should continue to undergo specialist review.

The high diagnostic performance observed despite brief training is consistent with emerging evidence that real-time AI guidance may reduce traditional technical barriers to echocardiographic acquisition. Mor-Avi et al. demonstrated that novice nurses and medical residents using AI guidance acquired images of sufficient quality in 81%–94% of examinations across standard views, with diagnostic concordance of 83%–96% relative to expert-acquired studies.^13^ In an accompanying editorial, Meucci and Delgado argued that real-time AI guidance may accelerate training by addressing one of echocardiography’s principal limitations: the dependence of image acquisition on operator skill and individualized probe manipulation.^14^ Our qualitative findings provide complementary real-world support for this mechanism. Nurse practitioners described AI as particularly valuable for probe guidance, image acquisition, and confidence building, while continuing to rely on clinical judgment and cardiologist review for abnormal or equivocal findings.

Similarly, Peck et al. showed that novice healthcare workers could obtain interpretable echocardiographic images suitable for RHD screening after less than five hours of AI-assisted training, particularly for mitral valve assessment.^15^ Whereas that study focused on image quality and interpretability, our study evaluated diagnostic accuracy using cardiologist overread as the reference standard and additionally explored implementation barriers and facilitators through qualitative interviews. Together, these studies suggest that AI-guided handheld echocardiography has progressed from demonstrating technical feasibility to supporting disease-specific screening programs in underserved populations.

The qualitative findings provide important context for interpreting these quantitative results. Participants consistently described competency as developing through repeated clinical practice rather than initial instruction alone, reinforcing previous implementation literature demonstrating that brief introductory training should be supplemented with additional supervised practice. Providers also consistently emphasized that AI functioned primarily as an image acquisition aid rather than an autonomous diagnostic system. Although AI improved confidence during image acquisition and reduced technical uncertainty, clinicians continued to rely on their own judgment and cardiologist consultation when evaluating abnormal or borderline findings. This observation mirrors previous AI-guided echocardiography studies and suggests that current AI technologies should be viewed as decision-support tools rather than replacements for specialist expertise.^13^ Furthermore, providers identified workflow, staffing, internet connectivity, patient flow, language support, and referral pathways as equally important determinants of successful implementation. These findings illustrate that successful deployment of AI-assisted screening depends not only on technological performance but also on thoughtful integration within existing healthcare systems.

Our findings also contribute to the broader evolution of RHD screening strategies. Longenecker previously argued that widespread implementation of echocardiographic screening would require simplified protocols, handheld devices, and non-expert operators to become logistically feasible and economically sustainable.^16^ The editorial also emphasized that additional implementation evidence was needed before community-based screening could be recommended outside research settings.^16^ Our study addresses one of these implementation gaps by demonstrating that nurse practitioners using AI-guided handheld echocardiography can accurately perform RHD screening in a rural community setting after a relatively brief training period. Although our study does not determine whether screening improves long-term clinical outcomes or reduces disease progression, it provides practical evidence supporting one of the essential components required for future population-level RHD screening programs.

This study has several strengths. To our knowledge, it is the first mixed-methods evaluation of AI-guided handheld echocardiography for RHD screening conducted in rural Nepal. The study was performed under real-world field conditions rather than a controlled research environment, increasing the applicability of the findings to community screening programs. By combining diagnostic accuracy with qualitative implementation data, we were able to evaluate not only whether the technology worked but also how it could be successfully integrated into rural healthcare delivery. Finally, the evaluation of AI-assisted task-shifting directly addresses a major workforce limitation affecting cardiovascular care in many low-and middle-income countries (LMICs).

Several limitations should also be acknowledged. The relatively small sample size and limited number of positive valve lesions reduced the precision of lesion-specific diagnostic estimates, especially for less common abnormalities. The absence of positive aortic stenosis cases prevented evaluation of device performance for this lesion. Participants were recruited through convenience sampling during community screening camps, which may limit generalizability to other populations and healthcare settings. Additionally, competency was assessed immediately following a brief training period, and long-term retention of screening skills was not evaluated. Larger multicenter studies with longer follow-up will be necessary to confirm these findings and evaluate reproducibility across different providers and populations.

The findings of this study have important implications for RHD control in Nepal and other LMICs. AI-guided handheld echocardiography performed by non-cardiologist providers offers a practical strategy for expanding access to screening while reducing dependence on specialist outreach services. Such an approach could facilitate more accurate estimation of national RHD burden, enable earlier initiation of secondary prophylaxis with benzathine penicillin G, and support implementation of the World Health Organization’s goals for RHD prevention and control. Given the device’s portability, relatively low cost, and ease of use, this intervention may also prove valuable for screening other structural heart diseases in underserved communities. Future research should focus on multicenter validation, long-term competency assessment, cost-effectiveness analyses, and integration of AI-assisted echocardiography into primary healthcare systems.

## Data Availability

All data is available upon request

## Acknowledgments

All listed authors contributed to this study.

## Supplemental material

- **Supplementary Appendix 1: Interview Guide**

- **Supplementary Appendix 2: Codebook**

## References

1. Gautam S, Dhungel S, KC B, Khan M, Shah S. Rheumatic Heart Disease In School Going Children: A Study From Lumbini Province, Nepal. J Univers Coll Med Sci. 2023;11(3):7–9.

2. Zhuang S, Guo D, Yu D. A mini review of the pathogenesis of acute rheumatic fever and rheumatic heart disease. Front Cell Infect Microbiol. 2025;15:1447149. doi:10.3389/fcimb.2025.1447149

3. Passos LSA, Nunes MCP, Aikawa E. Rheumatic Heart Valve Disease Pathophysiology and Underlying Mechanisms. Front Cardiovasc Med. 2021;7:612716. doi:10.3389/fcvm.2020.612716

4. Reményi B, Wilson N, Steer A, et al. World Heart Federation criteria for echocardiographic diagnosis of rheumatic heart disease—an evidence-based guideline. Nat Rev Cardiol. 2012;9(5):297–309. doi:10.1038/nrcardio.2012.7

5. Williamson J, Remenyi B, Francis J, et al. Echocardiographic Screening for Rheumatic Heart Disease: A Brief History and Implications for the Future. Heart Lung Circ. 2024 Jul;33(7):943–950. Doi 10.1016/j.hlc.2024.02.021

6. Rwebembera J, Marangou J, Mwita JC, et al. 2023 World Heart Federation guidelines for the echocardiographic diagnosis of rheumatic heart disease. Nat Rev Cardiol. 2024;21(4):250-263. doi:10.1038/s41569-023-00940-9

7. WHO guideline on the prevention and diagnosis of rheumatic fever and rheumatic heart disease. Geneva: WHO; 2024, https://www.who.int/publications/i/item/9789240100077

8. END RHD Campaign. Nepal Heart Foundation, Central Office. Accessed July 14, 2025. https://nepalheart.org.np/end-rhd-campaing-nepal/ [sic]

9. S L, K. Kaushal T, Subedi A, K. Sharma S. Underuse of Injectable Penicillin in Secondary Prophylaxis of Rheumatic Heart Disease in Nepal. Cardiol Vasc Res. 2020;4(5).

10. Rajbanshi BG, Pradhan S, Koirala B. Twenty-five years of Cardiac Surgery in Nepal: Trials, Tribulations and Triumph. Nepal Heart J. 2022;19(2):2. doi:10.3126/njh.v19i2.48936

11. Hosain N, Amin F, Maruf MF, et al. Global geographical discrepancy in numerical distribution of cardiovascular surgeries and human resource development in South Asia. JTCVS Open. 2022;11:192–199. doi:10.1016/j.xjon.2022.05.002

12. PEN-Plus Project – Kathmandu Institute of Child Health (KIOCH). Accessed November 22, 2025. https://kioch.org.np/projects/pen-plus-project/?utm

13. Mor-Avi V, Khandheria B, Klempfner R, et al. Real-Time Artificial Intelligence–Based Guidance of Echocardiographic Imaging by Novices: Image Quality and Suitability for Diagnostic Interpretation and Quantitative Analysis. Circ Cardiovasc Imaging. 2023;16(11). doi:10.1161/CIRCIMAGING.123.015569

14. Meucci MC, Delgado V. Artificial Intelligence to Speed Up Training in Echocardiography: The Next Frontier. Circ Cardiovasc Imaging. 2023;16(11). doi:10.1161/CIRCIMAGING.123.016148

15. Peck D, Rwebembera J, Nakagaayi D, et al. The Use of Artificial Intelligence Guidance for Rheumatic Heart Disease Screening by Novices. J Am Soc Echocardiogr. 2023;36(7):724–732. doi:10.1016/j.echo.2023.03.001

16. Longenecker CT. Echo Screening for Rheumatic Heart Disease: Are We There Yet? Circ Cardiovasc Imaging. 2019;12(2):e008818. doi:10.1161/CIRCIMAGING.118.008818

